# Stability of SARS-CoV-2 in different environmental conditions

**DOI:** 10.1101/2020.03.15.20036673

**Authors:** Alex W.H. Chin, Julie T.S. Chu, Mahen R.A. Perera, Kenrie P.Y. Hui, Hui-Ling Yen, Michael C.W. Chan, Malik Peiris, Leo L.M. Poon

## Abstract

Stability of SARS-CoV-2 in different environmental conditions.

## To the Editor

We previously reported the detection of SARS-CoV-2 in different clinical samples^1^. This virus can be detected on different surfaces in a contaminated site^2^. Here, we report the stability of SARS-CoV-2 in different environmental conditions.

We first determined the stability of SARS-CoV-2 at different temperatures. SARS-CoV-2 in virus transport medium (VTM; final concentration: ∼6.8 log TCID_50_/mL) was incubated for up to 14 days and then tested for its infectivity (Table A). The virus is highly stable at 4°C, but sensitive to heat. At 4°C, there was only ∼0.7-log unit reduction of infectious titre on Day 14. With the incubation temperature being increased to 70°C, the time for virus inactivation was reduced to 5 minutes.

**Table.** Stability of SARS-CoV-2 at different environmental conditions.

| Time | Virus titre (Log TCID<sub>50</sub>/mL) |  |  |  |  |  |  |  |  |  |
| --- | --- | --- | --- | --- | --- | --- | --- | --- | --- | --- |
|  | 4°C |  | 22°C |  | 37°C |  | 56°C |  | 70°C |  |
|  | Mean | ±SD | Mean | ±SD | Mean | ±SD | Mean | ±SD | Mean | ±SD |
| 1 min | N.D. | N.D. | 6.51 | 0.27 | N.D. | N.D. | 6.65 | 0.1 | 5.34 | 0.17 |
| 5 mins | N.D. | N.D. | 6.7 | 0.15 | N.D. | N.D. | 4.62 | 0.44 | U | - |
| 10 mins | N.D. | N.D. | 6.63 | 0.07 | N.D. | N.D. | 3.84 | 0.32 | U | - |
| 30 mins | 6.51 | 0.27 | 6.52 | 0.28 | 6.57 | 0.17 | U | - | U | - |
| 1 hr | 6.57 | 0.32 | 6.33 | 0.21 | 6.76 | 0.05 | U | - | U | - |
| 3 hrs | 6.66 | 0.16 | 6.68 | 0.46 | 6.36 | 0.19 | U | - | U | - |
| 6 hrs | 6.67 | 0.04 | 6.54 | 0.32 | 5.99 | 0.26 | U | - | U | - |
| 12 hrs | 6.58 | 0.21 | 6.23 | 0.05 | 5.28 | 0.23 | U | - | U | - |
| 1 day | 6.72 | 0.13 | 6.26 | 0.05 | 3.23 | 0.05 | U | - | U | - |
| 2 days | 6.42 | 0.37 | 5.83 | 0.28 | U | - | U | - | U | - |
| 4 days | 6.32 | 0.27 | 4.99 | 0.18 | U | - | U | - | U | - |
| 7 days | 6.65 | 0.05 | 3.48 | 0.24 | U | - | U | - | U | - |
| 14 days | 6.04 | 0.18 | U | - | U | - | U | - | U | - |

A) Temperature\* B) Surfaces\*
| Time | Virus titre (Log TCID<sub>50</sub>/ml) |  |  |  |  |  |  |  |  |  |
| --- | --- | --- | --- | --- | --- | --- | --- | --- | --- | --- |
|  | Paper |  | Tissue paper |  | Wood |  | Cloth |  | Glass |  |
|  | Mean | ±SD | Mean | ±SD | Mean | ±SD | Mean | ±SD | Mean | ±SD |
| 0 min | 4.76 | 0.10 | 5.48 | 0.10 | 5.66 | 0.39 | 4.84 | 0.17 | 5.83 | 0.04 |
| 30 mins | 2.18 | 0.05 | 2.19 | 0.17 | 3.84 | 0.39 | 2.84 | 0.24 | 5.81 | 0.27 |
| 3 hrs | U | - | U | - | 3.41 | 0.26 | 2.21" | - | 5.14 | 0.05 |
| 6 hrs | U | - | U | - | 2.47 | 0.23 | 2.25 | 0.08 | 5.06 | 0.31 |
| 1 day | U | - | U | - | 2.07" | - | 2.07" | - | 3.48 | 0.37 |
| 2 days | U | - | U | - | U | - | U | - | 2.44 | 0.19 |
| 4 days | U | - | U | - | U | - | U | - | U | - |
| 7 days | U | - | U | - | U | - | U | - | U | - |

| Time | Banknote |  | Stainless steel |  | Plastic |  | Mask, inner layer |  | Mask, outer layer |  |
| --- | --- | --- | --- | --- | --- | --- | --- | --- | --- | --- |
|  | Mean | ±SD | Mean | ±SD | Mean | ±SD | Mean | ±SD | Mean | ±SD |
| 0 min | 6.05 | 0.34 | 5.80 | 0.02 | 5.81 | 0.03 | 5.88 | 0.69 | 5.78 | 0.10 |
| 30 mins | 5.83 | 0.29 | 5.23 | 0.05 | 5.83 | 0.04 | 5.84 | 0.18 | 5.75 | 0.08 |
| 3 hrs | 4.77 | 0.07 | 5.09 | 0.04 | 5.33 | 0.22 | 5.24 | 0.08 | 5.11 | 0.29 |
| 6 hrs | 4.04 | 0.29 | 5.24 | 0.08 | 4.68 | 0.10 | 5.01 | 0.50 | 4.97 | 0.51 |
| 1 day | 3.29 | 0.60 | 4.85 | 0.20 | 3.89 | 0.33 | 4.21 | 0.08 | 4.73 | 0.05 |
| 2 days | 2.47 | 0.23 | 4.44 | 0.20 | 2.76 | 0.10 | 3.16 | 0.07 | 4.20 | 0.07 |
| 4 days | U | - | 3.26 | 0.10 | 2.27 | 0.09 | 2.47 | 0.28 | 3.71 | 0.50 |
| 7 days | U | - | U | - | U | - | U | - | 2.79 | 0.46 |

C) Disinfectants\*
| Disinfectant<br>(Working concentration) | Virus titre (Log TCID<sub>50</sub>/mL) |  |  |
| --- | --- | --- | --- |
|  | 5 mins | 15 mins | 30 mins |
| Household bleach (1:49) | U | U | U |
| Household bleach (1:99) | U | U | U |
| Hand soap solution (1:49) | 3.6" | U | U |
| Ethanol (70%) | U | U | U |
| Povidone-iodine (7.5%) | U | U | U |
| Chloroxylonol (0.05%) | U | U | U |
| Chlorhexidine (0.05%) | U | U | U |
| Benzalkonium chloride (0.1%) | U | U | U |

D) pH\*
| pH<br>(60 mins) | Virus titre<br>(Log TCID<sub>50</sub>/mL) |  |
| --- | --- | --- |
|  | Mean | ±SD |
| 3 | 5.55 | 0.25 |
| 4 | 5.67 | 0.36 |
| 5 | 5.73 | 0.04 |
| 6 | 5.75 | 0.08 |
| 7 | 5.58 | 0.22 |
| 8 | 5.70 | 0.14 |
| 9 | 5.54 | 0.44 |
| 10 | 5.51 | 0.11 |

\* All the virus titres were titrated using Vero-E6 cells. All experimental studies were done in three independent triplicates. Detection limit of a typical TCID<sub>50</sub> assay is 100 TCID<sub>50</sub>/mL, except reactions containing hand soap/chloroxylonol (detection
limit: $10^3$ TCID<sub>50</sub>/mL) or reactions containing povidone-iodine/cholorhexidin/benzalkonium chloride; detection limit: $10^4$ TCID<sub>50</sub>/mL) because of their cytotoxic effects. N.D.: not done, U: undetectable.

We further investigated the stability of this virus on different surfaces. In brief, a 5-µL droplet of virus culture (∼7.8 Log unit of TCID_50_/mL) was pipetted on a surface (Table B; ∼1cm^2^ per piece) and left at room temperature (22°C; Relative humidity: ∼65%). The inoculated objects retrieved at desired time points were immediately soaked with 200 µL of VTM for 30 minutes to elute the virus. No infectious virus could be recovered from printing and tissue papers after a 3-hour incubation, whereas no infectious virus could be detected from treated wood and cloth on Day 2. By contrast, SARS-CoV-2 was more stable on smooth surfaces. No infectious virus could be detected from treated smooth surfaces on Day 4 (glass and banknote) or Day 7 (stainless steel and plastic). Strikingly, a significant level of infectious virus could still be detected on the outer layer of a surgical mask on Day 7 (∼0.1% of the original inoculum). Interestingly, a biphasic decay of infectious SARS-CoV-2 could be found from samples recovered from these smooth surfaces (Appendix). Representative negative samples were tested positive by RT-PCR^3^ (N=39; data not shown), demonstrating that non-infectious viruses could be recovered by the eluents.

We also tested the virucidal effects of disinfectants by adding 15 µL of SARS-CoV-2 culture (∼7.8 Log unit of TCID_50_/mL) to 135 µL of various disinfectants at working concentration (Table C). With the exception of a 5-min incubation with hand soap, no infectious virus could be detected after a 5-minute incubation at room temperature. In addition, we also found that SARS-CoV-2 is extremely stable in a wide-range of pH values at room temperature (pH3-10; Table D)

Overall, SARS-CoV-2 can be highly stable in a favourable environment^4^, but it is also susceptible to standard disinfection methods.

We declare that we have no competing interests. This work was supported by NIADI, NIH (USA) (contract HHSN272201400006C). LLMP was supported by Croucher Foundation.

## Data Availability

All data will be available upon request

## Notes

### Competing Interest Statement

The authors have declared no competing interest.

